# Gender differences in the perception of leptospirosis severity, behaviours, and *Leptospira* exposure risk in urban Brazil: a cross-sectional study

**DOI:** 10.1101/2024.04.28.24306445

**Authors:** Ellie A. Delight, Diogo César de Carvalho Santiago, Fabiana Almerinda G. Palma, Daiana de Oliveira, Fábio Neves Souza, Juliet Oliveira Santana, Arata Hidano, Yeimi Alexandra Alzate López, Mitermayer Galvão G. Reis, Albert I. Ko, Akanksha A. Marphatia, Cleber Cremonense, Federico Costa, Max T. Eyre

**Affiliations:** London School of Hygiene & Tropical Medicine, London, UK; Federal University of Bahia, Salvador, Collective Health Institute, Salvador, Bahia, Brazil; Instituto Gonçalo Moniz, Fundação Oswaldo Cruz, Ministério da Saúde, Salvador, Bahia, Brazil; Department of Epidemiology of Microbial Diseases, Yale School of Public Health, New Haven, CT, USA

**Keywords:** disadvantaged urban communities, informal settlement, gender, sex-disaggregated, behaviour, disease risk perceptions, leptospirosis, flooding

## Abstract

**Background:** Vulnerability to climate hazards and infectious diseases are not gender-neutral, meaning that men, women, boys, girls, and other gender identities experience different health risks. Leptospirosis, a zoonotic climate sensitive infectious disease, is commonly transmitted to humans via contact with animals and the environment, particularly soil and flood water. Gender differences in leptospiral infection risk are reported globally, with men consistently found to be at higher risk than women. However, the drivers of this difference in risk are poorly understood. Previous studies suggest that the interplay of knowledge, perceptions, and behaviours may shape differential infection risk among genders.

**Methodology/Principal Findings:** To examine gender differences in *Leptospira* exposure risk we conducted a cross-sectional serosurvey among adult participants (n = 761) in four urban, marginalised, informal settlements in the city of Salvador, Brazil. We found that seroprevalence was 14.6% and 9.4% across men and women respectively. We then applied causal inference methodology to a two-part sex-disaggregated analysis to investigate: 1) the association of perceptions and behaviours with *Leptospira* seropositivity and 2) the association of perceptions with behaviours. We found that men who perceived leptospirosis as extremely serious had lower odds of seropositivity, walking through sewage water, or walking barefoot, suggesting an important link between perceptions, behaviours, and exposure risk. These associations were not found in women, and these behaviours were not associated with seropositivity in men or women.

**Conclusions:** Our results highlight perceived severity of disease as a potential driver of behaviour in men, and perceptions of disease may be an important target for health education programs. Furthermore, our study identifies evidence gaps in the understanding of infection risks in women. As the first sex-disaggregated study investigating *Leptospira* infection risks, we advocate for a gendered lens in future studies to further understand risks specific to different gender identities.

**Author summary:** Leptospirosis is a wide-spread zoonotic pathogen commonly spread from rodents to humans in urban informal settlements vulnerable to flooding in Salvador, Brazil. Gendered cultural norms shape perceptions of leptospirosis, behaviour, and subsequent exposure at the human-animal-environment interface. Despite this, there is limited research investigating gender-determined infection risks. Therefore, our study investigated associations of risk factors for *Leptospira* seropositivity in men and women. We identified perceived severity of leptospirosis, high-risk occupations, and age as significant risk factors in men. We also investigated perceived severity as a driver of risk by estimating the association of perceived severity with behaviours. We found that greater perceived severity was associated with reduced odds of walking through sewage water and walking barefoot outside of the home in men, however this behaviour was not significantly associated with reduced odds of leptospirosis. Our results suggest evidence gaps in the understanding of transmission routes in women. As the first sex-disaggregated study investigating gender-determined *Leptospira* transmission routes, our findings illustrate the importance of gendered behaviours, perceptions, and risk as drivers of leptospirosis, and highlight the need for further research to understand exposures more prevalent in women. We advocate for a greater focus on gender to help unravel gender-determined infection risks.

## Introduction

As climate change progresses, climate-related hazards such as flooding are becoming more frequent and severe, posing significant risks to human health. A recent systematic review revealed that 58% of human infectious diseases are exacerbated by climatic hazards, identified as climate-sensitive infectious diseases (CSIDs) (1). The review highlighted that CSIDs are transmitted to humans through 1,006 unique pathways sensitive to climate hazards, with 121 of these pathways attributed to flooding. These diseases are predicted to disproportionately affect urban informal settlements which are vulnerable to the effects of flooding events due to inadequate provision of drainage systems and other basic urban services.

Vulnerability to both flooding and CSIDs is not gender-neutral, meaning that men, women, boys, girls, and other gender identities living in the same environments experience different health risks (2–5). For example, men can face greater risk of exposure to CSID pathogens through contact with the environment and animals attributed to outdoor occupations, while women can face greater socioeconomic effects of CSIDs due to heightened job insecurity (6–8). Additionally, adverse health outcomes in women are accentuated during flooding due to limited mobility or access to services, and traditional caring roles, rendering them less climate resilience (9–11).

Flooding is an important driver of leptospirosis, a zoonotic neglected CSID caused by pathogenic bacteria of the genus *Leptospira* (1,4,8,12,13). In urban informal settlements, leptospires are shed into the environment in the urine of the infected rat reservoir, where they can survive for days or weeks (14). The persistence of leptospires makes the environment an important reservoir and source of infection for humans through contact with soil, mud, and water. Flooding exacerbates this problem by dispersing leptospires across the environment and bringing humans directly into contact with contaminated flood water, increasing the risk of spillover transmission at the human-animal-environment interface (4,8,15–17).

Leptospirosis is more prevalent in men, who suffer from a higher estimated global health burden and are reported as being at greater risk of *Leptospira* infection across a wide range of geographical contexts (13). This is the case in climate-vulnerable urban informal settlements in Brazil where men are reported to have over twice the risk of *Leptospira* infection than women (4,8,18,19). While the mechanisms driving this gender difference in infection rates are poorly understood (4,14,17,18), they have largely been attributed to exposures that are more prevalent in men, and there is no evidence to suggest that they are caused by physiological differences between sexes (8). Whilst previous studies have investigated the odds of one sex having a higher risk of exposures or *Leptospira* seropositivity compared to another sex, none have examined whether different pathways shape this risk for the two sexes (or different genders), by for example, conducting sex-disaggregated analysis of factors shaping risk. Notably, the disease burden in women should not be underestimated; misdiagnosis with pregnancy-related conditions in women is common and can result in maternal and foetal death (20), and active surveillance in Salvador from 2003 – 2005 reported that while the majority of leptospirosis-associated Severe Pulmonary Haemorrhagic Syndrome (SPHS) patients were male (70%), women had almost three times the odds of developing SPHS compared to men (OR: 2.87, 95%CI: 1.36, 5.98) (21).

While research employing a gender perspective to the study of leptospirosis remains scarce, several studies in the city of Salvador, Brazil, have suggested that behaviour may be an important determinant of the gender-specific risks associated with the disease (8,22,23). For example, a previous study used GPS to monitor residents’ mobility and found that men covered a larger area than women over a 24-hour period, as did infected male participants when compared with uninfected males (23). However, exposure to environmental sources of leptospirosis transmission did not vary by gender, although this may have been due to challenges in accurately measuring environmental exposure (23). A knowledge, attitudes, and practices (KAP) study, also conducted in Salvador, found that men had lower leptospirosis knowledge scores compared to women, which was associated with reduced protective hygiene-related behaviours when in contact with the environment, such as wearing gloves when handling waste (8). Upon adjusting for KAP, male gender was no longer a risk factor for *Leptospira* infection, suggesting that the higher rates in men could be attributed to differences in knowledge and attitudes, which shape behaviours. This finding aligns to the Rational Model Theory (24), that behaviour is determined by knowledge and perceptions of disease (25–27). Collectively, these findings suggest that a complex interplay between social norms, knowledge, perceptions, and behaviours may result in varying levels of exposure to the contaminated environment, and consequently *Leptospira* infection risk, among genders.

The use of causal inference methodology and application of sex-disaggregated frameworks are important for effective study of gender-determined health risk. Causal inference methodology, utilising directed acyclic graphs (DAGs), enables researchers to map out causal systems and assumptions, offering a visual representation of complex relationships, including those related to gender-determined infection risk for *Leptospira* (28). Disaggregating epidemiological analyses by sex and gender, as recommended in the WHO toolkit for incorporating intersectional gender analysis into research on infectious diseases of poverty, allows for a gender-sensitive focused examination of infection risks (29). Investigating gender-determined disease infection risks can generate evidence that supports tailoring of interventions against leptospirosis for the specific needs of all genders. Additionally, it provides a foundation for further intersectional analyses in future studies to examine how other inequalities, such as race and socioeconomic status, intersect with gendered risks.

Understanding the underlying drivers of exposure to the environment for men and women is increasingly important as climate change continues to expose marginalised individuals to CSIDs and embed gendered inequalities. The aim of this study was to understand how *Leptospira* infection risk varies by gender among men and women living in four informal communities in Salvador, Brazil, by considering the role of perceptions and behaviours within a sex-disaggregated framework. First, we investigate the relationship between sociodemographic factors, perceptions, and behaviours and *Leptospira* infection in men and women. Then, we investigate how perceived severity of leptospirosis impacts behaviours in men and women. Therefore, this study also aims to provide a methodological example of sex-disaggregated analyses to advocate for gender-sensitive methodologies in CSID research.

## Materials and methods

### Study design

#### Study area

The study was conducted in four informal settlement communities in the city of Salvador, North-eastern Brazil: Nova Sussuarana (NS), Arenoso, Jardim Santo Inácio/Mata Escura (JSI/ME), and Calabetão (Figure 1A-B). The communities were identified as high-risk for *Leptospira* by surveillance data collected between 1996-2018 (30). All communities are characterised as low-income, urban informal settlements, with poor sanitation, drainage and waste management systems. The study site in each community was defined as an area within 40 metres of an open sewer, identified through a previously conducted census (30).

**Figure 1:**
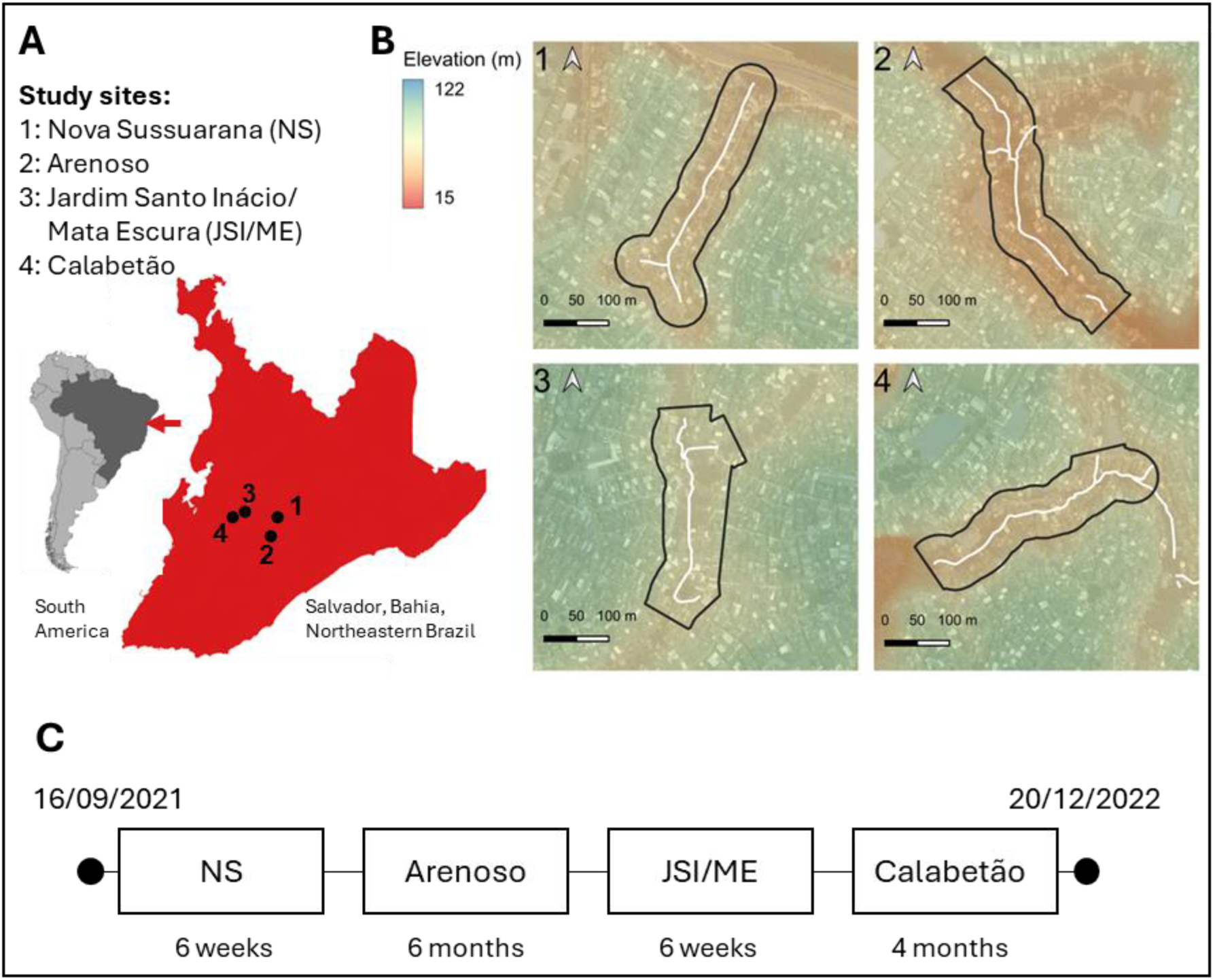
Study site and timeline. A) Position of study sites within the city of Salvador, Brazil shown as numbered points; B) Study sites shown as polygons with elevation and the position of open sewer indicated with white line; C) Cross-sectional serosurvey timeline.

#### Questionnaires and serosurveys

We conducted a cross-sectional study between September 2021 – December 2022 (Figure 1C). After an initial census, individuals were invited to be included in the study if they: had lived in ground floor households in the study area for at least 6 months, had slept at the property for at least 3 nights in the previous week, and were aged 18 years or older (30). Written informed consent was obtained from each participating individual.

Serosurveys were conducted within the study sites over a 15-month period and the duration of data collection in sites differed due to adverse weather and safety challenges. Data was collected during mornings from Monday – Sunday in attempt to include both employed and unemployed participants and mitigate selection bias. Trained phlebotomists visited participant homes, collected blood samples, and conducted a modified version of a standardised that has been validated previously (4,19). Houses were revisited up to five times in participants were not present at the time of the previous visit. Information about sociodemographic indicators (age, sex, race, completion of primary education, employment status, occupation), perceptions, and behaviours that relate to exposure was collected. Sex and race were self-reported. Employment status was defined as whether a participant had been employed during the previous week. High-risk occupations were defined as construction, street sweepers or vendors, recycling collectors, or cleaning services. Most behaviours (walking through flood water, sewage water, mud, or walking barefoot outside of the home) were characterised as the frequency at which an individual had performed a given behaviour in the previous six months (never, rarely, sometimes, often, always). Participants were also asked if they were able wear boots during flooding in the previous six months (owned boots, could borrow boots, or couldn’t access boots). Perceived severity was ranked on a five-point scale from not serious to extremely serious. Data was collected and securely stored using Research Electronic Data Capture (REDCap) electronic data collection tools hosted at Oswaldo Cruz Foundation (Fiocruz), Bahia (31,32).

Microscopic agglutination test (MAT), the reference assay of sero-diagnosis of leptospirosis, was used to measure titres of agglutinating *Leptospira* antibodies (33). Serological samples were tested for *Leptospira* strains: *L. kirschneri* serogroups Cynopteri serovar Cynopteri strain 3522C and Grippothyphosa serovar Grippothyphosa strain Duyster; *L. interrogans* serogroups Canicola serovar Canicola strain H. Utrecht IV and Autumnalis serovar Autumnalis strain Akiyami A; and *L. borgpetersenii* serogroup Ballum serovar Ballum strain MUS 127. The panel also included two local clinical isolates: *L. interrogans* serogroup Icterohaemorrhagiae serovar Copenhageni strain Fiocruz L1-130 and *L. santarosai* serogroup Shermani strain LV3954. Individuals were classified as seropositive if their serological sample returned an MAT titre of ≥1:50 for any serogroup. Laboratory testing was conducted in the Laboratory Pathology and Molecular Biology at Fiocruz, Salvador.

### Causal inference framework

A causal inference approach was used to investigate how *Leptospira* infection risk varies by gender with language used herein following that outlined by Tennant at al., 2021(28). A DAG was used to map the causal hypotheses underlying this study and to identify a sufficient set of adjustment variables in subsequent regression models (28). The DAG was created on Dagitty (34) and a simplified version is shown in Figure 2 (the full version can be found at https://dagitty.net/dags.html?id=XxPTXytr or Supplementary Information 1). For the development of the DAG, causal relationships between exposures and seropositivity were considered using plausible causal assumptions and assessed under temporality and biological of the Bradford Hill criterion (35). Our key causal assumption, as mapped in Figure 2, is that gender is a distal causal factor that influences risk of *Leptospira spp.* seropositivity. We hypothesise that gender acts directly on seropositivity, through unmeasured variables currently unknown, and indirectly through perceptions of disease, behaviour, and household environments.

**Figure 2:**
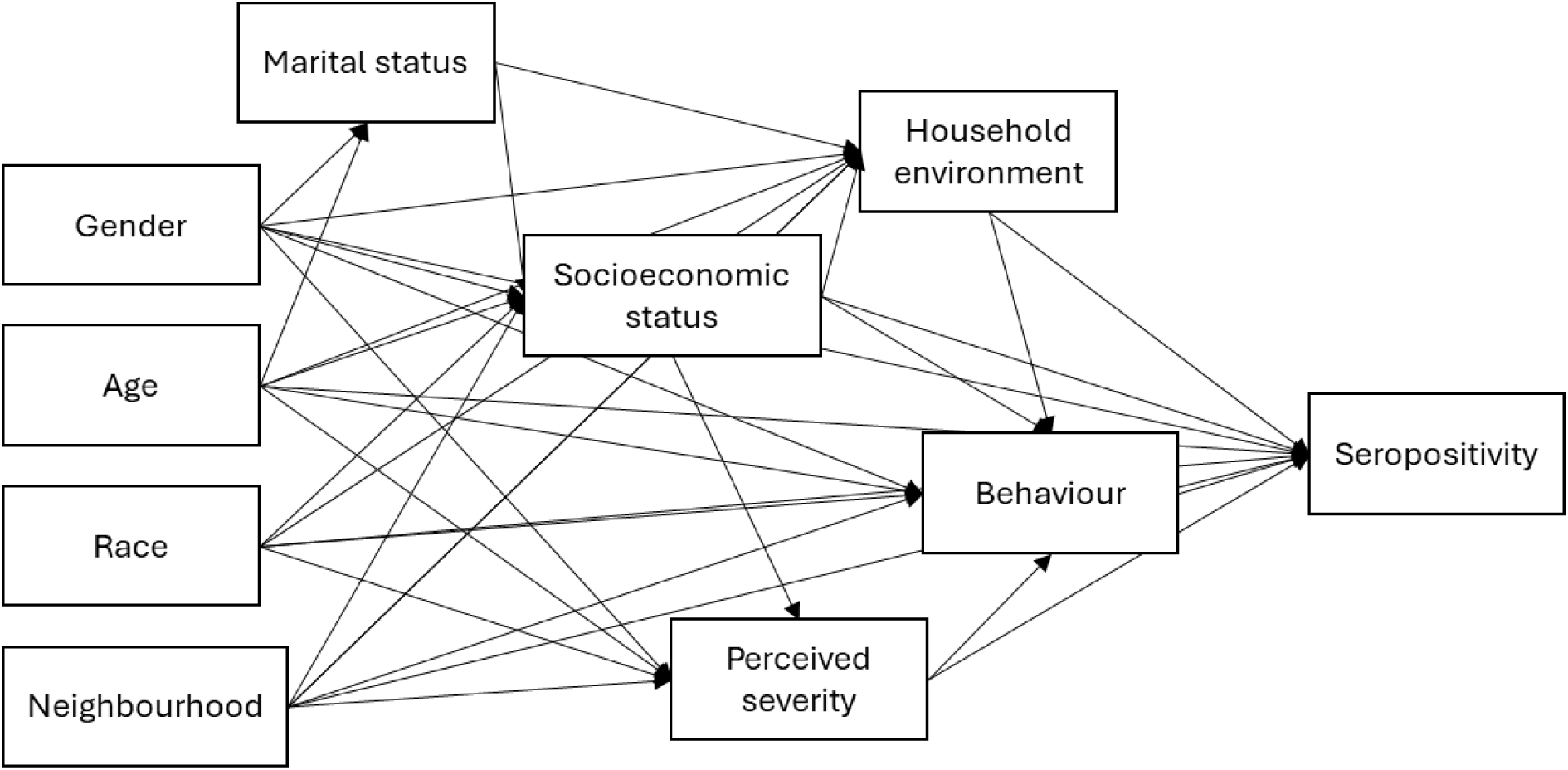
Simplified DAG, where direction of causality is indicated by arrows. Socioeconomic status: household food insecurity, education, employment, occupation; Household environment: flooding in household, living within 10m of open sewers of waste accumulation, and count of rats sighted around the household; Behaviour: Walking through sewage water, mud, or floodwater, ownership of boots, or walking barefoot.

### Statistical Analysis

Study participation was assessed aligned with the STROBE (STrengthening the Reporting of OBservational studies in Epidemiology) statement for cross-sectional studies (36). Chi-squared tests were used to compare non-participation rates across sociodemographic indicators available among both those that agreed and disagreed to participate (neighbourhood, age, and sex).

A sex-disaggregated analysis was conducted to identify gender-determined risk factors for *Leptospira* infection. The analysis was conducted in two parts (Figure 3), to estimate the total causal effect of: 1) sociodemographic, perception, and behavioural variables on *Leptospira* seropositivity, and 2) perceptions on behaviours. Due to data availability, our study investigates the genders of men and women and uses these terms interchangeably with male and female based on the UNICEF definition of gender as “differences by sex and the unique needs of males and females [which] reflect differences by gender, the socially and culturally constructed roles, responsibilities, and expectations of men and women, and boys and girls” (37).

**Figure 3:**
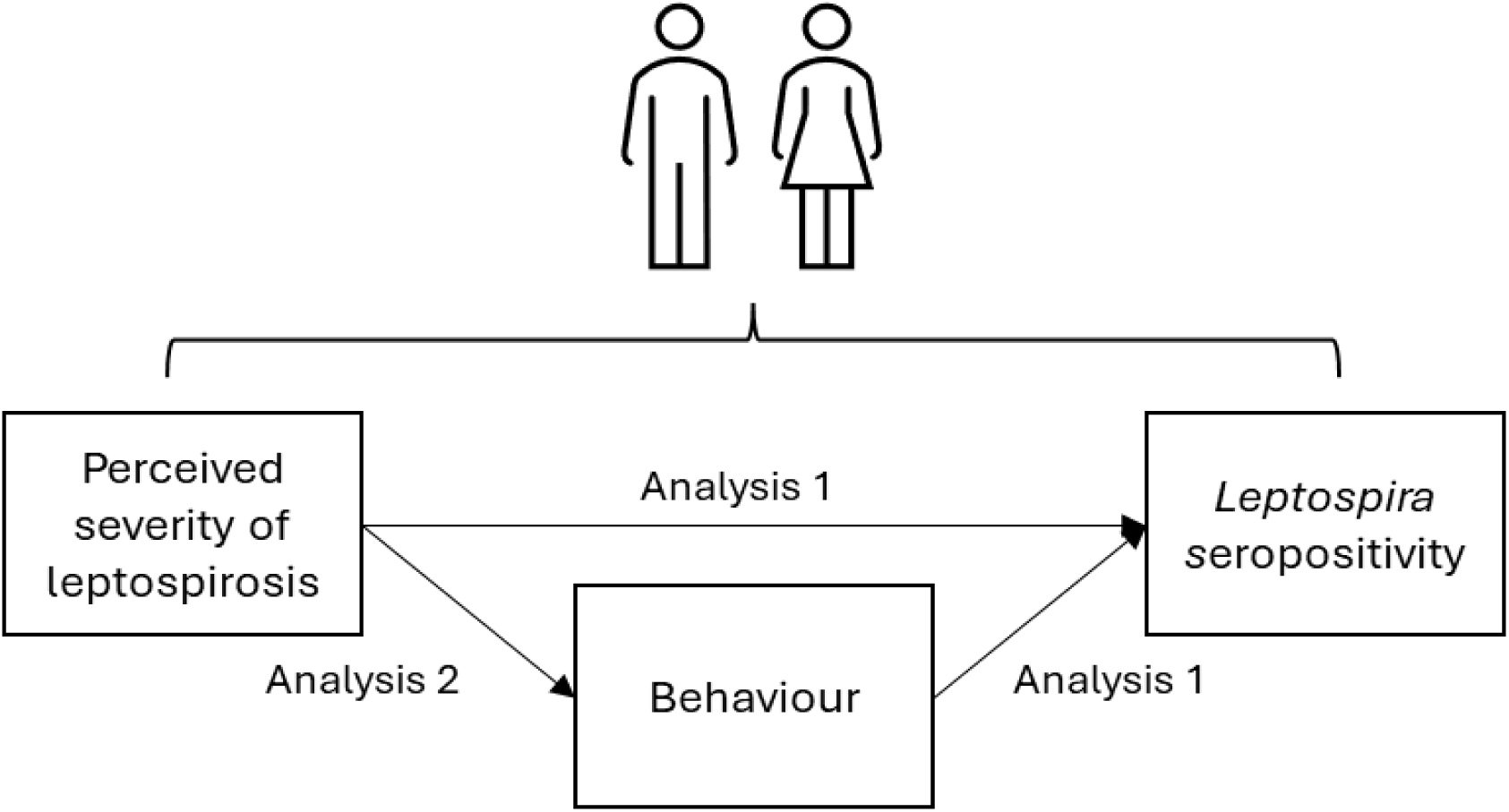
Hypothesised causal pathway of how perceived severity of leptospirosis causes Leptospira seropositivity in men and women. Analysis 1: Seropositivity risk actor analysis; Analysis 2: Perceived severity as a determinant of high-risk behaviours.

#### Analysis 1: Seropositivity risk factor analysis

In Analysis 1 we estimated the total causal effect of each sociodemographic, perception, and behaviour variable (exposures) on *Leptospira* seropositivity (outcome) in men and women.

##### Descriptive

Descriptive cross tabulations were used to describe the prevalence of exposures across seropositivity by gender.

##### Generalised additive models (GAMs) and regression analyses

GAMs were used to assess linearity and identify the functional form of the univariable relationship between age, a continuous explanatory variable, and seropositivity (38,39).

Univariable binomial generalised logistic regression mixed-effect models (GLMMs) were used to estimate the crude total causal effect of each exposure on seropositivity, including a household-level random effect to account for household clustering.

Multivariable GLMMs were used to estimate the adjusted total causal effect of each exposure on seropositivity by adjusting for the sufficient adjustment sets identified in the DAG (34). Each adjustment set was tested for multicollinearity using Cramer’s V Correlation test, and no adjustment sets showed evidence of collinearity between categorical variables (Cramer’s V coefficient < 0.08) (40). Mediator-outcome confounders (defined as variables that are confounders for the relationship between a given mediator and outcome pair, these are equivalent to competing exposures in Figure 2) were added to the minimal adjustment sets to improve estimate precision (28). Mediator-outcome confounders differed by model but were limited to age, race, season, and community. For both univariable and multivariable models, the effect of each exposure on seropositivity was estimated for the following sex-combined and sex-disaggregated populations: i) male and female; ii) male-restricted; iii) female-restricted. This resulted in three models per exposure. Only model coefficients for exposure variables were reported to avoid “Table 2 fallacy”, whereby coefficients of confounders are displayed and mistakenly interpreted (34,39).

Finally, the strength of evidence supporting sex as an effect modifier for the relationship between each exposure and outcome pair was investigated by including an interaction term in the multivariable model.

#### Analysis 2: Perceived severity as a determinant of high-risk behaviours

In analysis 2 we estimated the total causal effect of perceived severity of leptospirosis (exposure) on the risk of performing high-risk behaviours (outcome). These behaviours related to personal hygiene practices and the frequency of contact with known sources of environmental risk: being able to wear boots during flooding, walking through flood water, sewage water, and mud, walking barefoot outside of the home.

The same modelling approach was followed for these exposure-outcome pairs as for Analysis 1. Categorical behaviour variables with multiple levels were recategorised for inclusion into binomial models as response variables. Behavioural variables were dichotomised into “rarely” (inclusive of “never” and “rarely” observations) and “frequent” (inclusive of “sometimes”, “often”, and “always” observations). The ability to wear boots during flooding was dichotomised “yes” (inclusive of “yes, can borrow”, and “yes”) or “no” (inclusive of “no”). Most participants ranked leptospirosis as an extremely serious disease. Therefore, to account for the small number of observations across lower rankings, the perceived severity variable was dichotomised into extremely serious (inclusive of “extremely serious” only) and less serious (inclusive of “not serious”, “a little serious”, “serious” and “very serious”).

Age was modelled based on the functional forms identified by the GAMs in Supplementary Information 3c. Functional forms included linear forms, or linear piecewise splines with knots at identified ages.

Data were cleaned and analysed in R version 4.3.1 (41). Various R packages were used including ‘lme4’, ‘splines’, and ‘qgam’ (19,42–46).

### Ethics

Ethical approval for this study was obtained from the ethics committee at the Collective Health Institute, Federal University of Bahia (CEP/ISC/UFBA) under number CAEE 32361820.7.0000.5030, and the national research ethics committee (CONEP) linked to the Brazilian Ministry of Health under approval number 4.235.251. All participants involved in the study provided written informed consent before data collection.

## Results

### Study overview

#### Study participation

Across the four communities, we identified 688 households that were eligible using a baseline community census and household visits, 657 of which contained individuals aged ≥ 18 years and were eligible for inclusion in this analysis (Figure 4) (30). Of the 657 households, 481 (73%) head-of-households gave consent to participate, which permitted 916 residents eligible to participate. Of these 916 residents, 147 (16%) declined to participate and 8 (1%) were excluded from this analysis due to undetermined serostatus. Non-participation rates were similar across community (12%, 20%, 16%, and 16% in NS, Arenoso, JSI/ME, and Calabetão respectively, χ^2^ = 5.4, *df* = 3, *p* = 0.1). Individuals who declined to participate were younger on average than those who participated (38 vs. 42 years respectively, t-value = −2.6, *df* = 204.7, *p* = 0.009) and were significantly more likely to be male than female (70% vs. 37% respectively, χ^2^ = 52.4, *df* = 1, *p* < 0.001).

**Figure 4:**
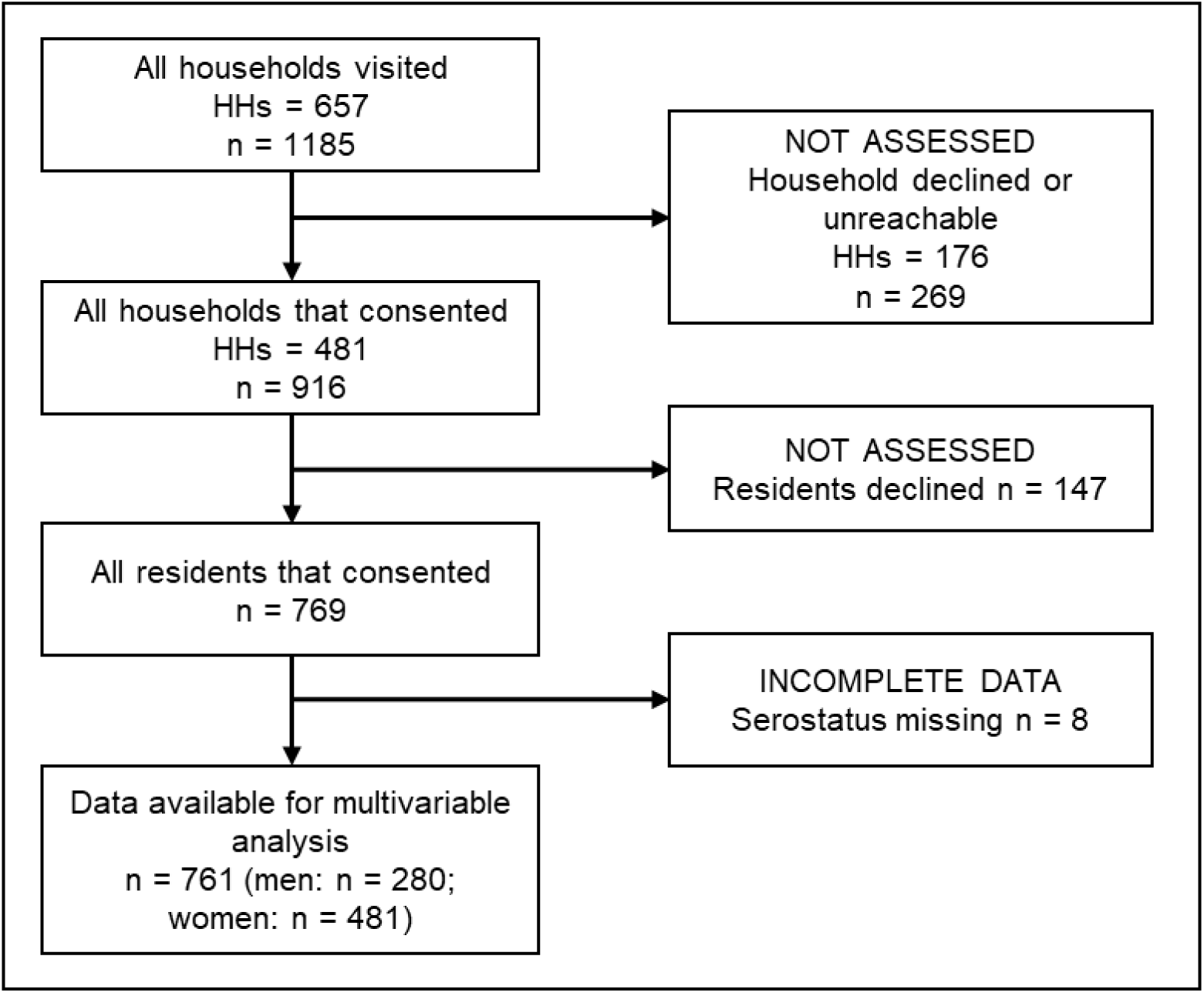
Study participant flow chart aligned with the STROBE (Strengthening the Reporting of OBservational Studies in Epidemiology) statement (36).

#### Study population demographics, perceptions, and behaviours

Of the 761 participants included in the analysis, 481 (63%) were female, the age ranged from 18 – 86 years, and the age distribution was similar across men and women (Table 1). The racial distribution was similar across genders, and most participants identified as Pardo (43%), indicating mixed ethnic backgrounds, or black (51%). Unemployment was higher in women than men (58% vs. 34% respectively, χ^2^ = 40.0, *df* = 1, *p* <0.001).

**Table 1:** Sex-disaggregated study population demographics, perceptions, and behaviours.

|  | N | Overall, N =<br>761<sup>1</sup> (%) | Female, N =<br>481<sup>1</sup> (%) | Male, N =<br>280<sup>1</sup> (%) | p-value* |
| --- | --- | --- | --- | --- | --- |
| **Sociodemographic** |  |  |  |  |  |
| Age | 761 |  |  |  | 0.6 |
| 18-30 |  | 202 (27) | 124 (26) | 78 (28) |  |
| 31-45 |  | 270 (35) | 169 (35) | 101 (36) |  |
| 46-60 |  | 191 (25) | 120 (25) | 71 (25) |  |
| >60 |  | 98 (13) | 68 (14) | 30 (11) |  |
| Race | 761 |  |  |  | 0.7 |
| Black |  | 386 (51) | 241 (50) | 145 (52) |  |
| Other |  | 48 (6) | 33 (7) | 15 (5) |  |
| Pardo |  | 327 (43) | 207 (43) | 120 (43) |  |
| Incomplete primary education | 761 | 600 (79) | 379 (79) | 221 (79) | >0.9 |
| Employment | 759 |  |  |  | <0.001 |
| Unemployed | 370 | 370 (49) | 276 (58) | 94 (34) |  |
| Formal | 760 | 159 (21) | 65 (14) | 94 (34) |  |
| Informal | 389 | 230 (30) | 139 (29) | 91 (33) |  |
| High-risk occupation<br>(among employed) | 389 | 95 (24) | 49 (24) | 46 (25) | 0.8 |
| **Serostatus** |  |  |  |  |  |
| Seropositive | 761 | 86 (11) | 45 (9.4) | 41 (15) | 0.03 |
| **Perceptions of leptospirosis** |  |  |  |  |  |
| Leptospirosis severity<br>perceived as "extremely<br>serious" | 746 | 649 (87) | 411 (87) | 238 (88) | 0.8 |
| **Behaviours conducted in last 6 months** |  |  |  |  |  |
| Walked through floodwater | 746 | 186 (25) | 126 (27) | 60 (22) | 0.2 |
| Walked through sewage<br>water | 747 | 161 (22) | 100 (21) | 61 (22) | 0.7 |
| Could wear boots during<br>flooding | 748 | 222 (30) | 86 (18) | 136 (50) | <0.001 |
| Walked barefoot outside of<br>home | 747 | 174 (23) | 108 (23) | 66 (24) | 0.7 |
| Walked through mud | 748 | 197 (26) | 120 (25) | 77 (28) | 0.4 |
| *Pearson's Chi-squared test |  |  |  |  |  |

Seroprevalence for *Leptospira*-specific antibodies was greater in men than women (15% vs. 9% respectively, χ^2^ = 4.9, *df* = 1, *p* = 0.03). Out of the 86 seropositive individuals, the majority were seropositive for Icterohaemorrhagiae strain Fiocruz L1-130 (69%) or Cynopteri strain 3522C (19%) serogroup. Leptospirosis was perceived as an extremely serious disease by a large proportion of both men and women (88% vs. 87% respectively, χ^2^ = 0.1, *df* = 1, *p* = 0.8). The proportions of men and women frequently conducting high-risk behaviours (walking through sewage water, flood water, mud, or walking barefoot outside) were similar. However, a significantly larger proportion of women lacked access to boots to wear during flooding than men (82% vs. 50% respectively, χ^2^ = 83.1, *df* = 1, *p* <0.001).

### Analysis 1: Seropositivity risk factor analysis

#### Descriptive

Seroprevalence differed by perceived severity of leptospirosis and age group in both men and women (Figure 5). Seroprevalence increased across age groups for both men and women although the percentage increase was greater in men than women (26.9% increase across 18-30 to >60 age groups in men, vs. 8.2% increase across 18-30 and >60 age groups in women). Seroprevalence was also lower among both men and women who perceived leptospirosis as an extremely serious disease than those who didn’t (13.0% vs. 26.5% in men and 8.8% vs. 14.3% respectively in women). The sex-disaggregated descriptive analysis of seroprevalence across all risk-factors investigated in this analysis is included in Supplementary Figure 2a.

**Figure 5:**
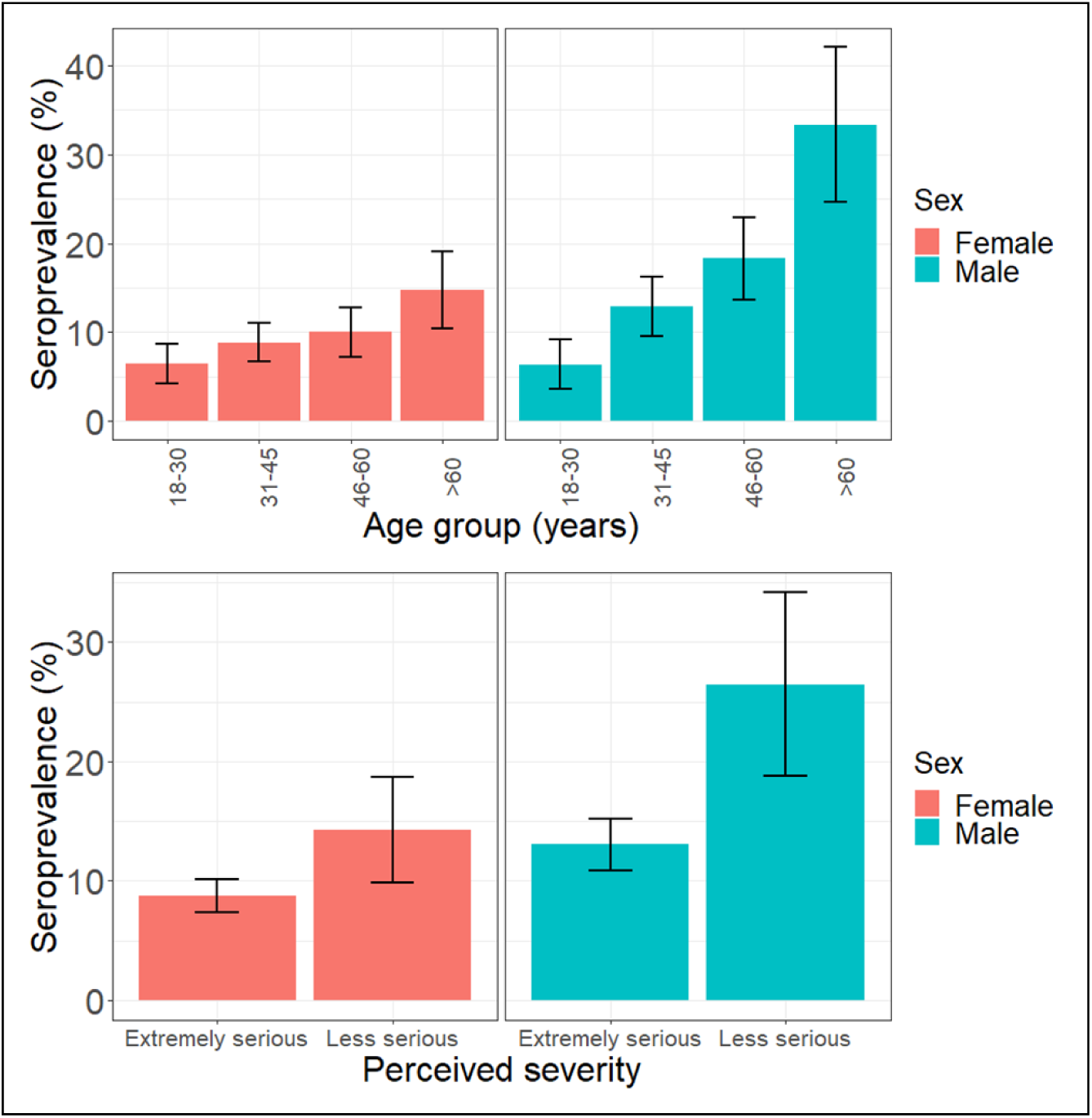
Sex-disaggregated seroprevalence across age group and perceived severity.

#### Regression analyses

Uni- and multivariable analyses were used to estimate the total causal effect of each exposure on seropositivity. The univariable analysis is included in Supplementary Information 2b. In each of the multivariable regression models (Table 2), age was modelled as a continuous variable based on the functional form identified in the GAM analysis (Supplementary Information 2b). In the multivariable analysis, men had 1.76 (95%CI 1.11, 2.79) times the odds of *Leptospira* seropositivity than women.

Older age was associated with increased odds of seropositivity in the male-restricted and combined models, but not in female-restricted model. In the male-restricted model, each year of age in men was associated with 1.04 (95%CI 1.01, 1.06) times the odds of seropositivity. In the combined model, each year of age was associated with 1.03 (95%CI 1.01, 1.04) times the odds of seropositivity. A similar estimate was found in the female-restricted model, as each year of age was associated with 1.02 (95%CI 0.99, 1.05) times the odds of seropositivity, however the association was not significant at the 5% level. A test for interaction showed no evidence that the association of age with seropositivity differed significantly between genders (*p*-value of interaction = 0.2).

Greater perceived severity was associated with reduced odds of seropositivity in the male-restricted and combined models, but not in female-restricted model. In the male-restricted model, men who perceived leptospirosis as extremely serious had 0.38 (95%CI 0.15, 0.99) times the odds of being seropositive than men who perceived leptospirosis as less serious. In the combined model, those that perceived leptospirosis as extremely serious had 0.55 (95%CI 0.30, 0.99) times the odds of being seropositive than those that perceived leptospirosis as less serious. A similar estimate was found in the female-restricted model, with women that perceived leptospirosis as extremely serious having 0.54 (95%CI 0.18, 1.60) times the odds of being seropositive than women who perceived leptospirosis as less serious, however the association was not significant at the 5% level. The associations in the male- and female-restricted models are visualised in Figure 6A. A test for interaction showed no evidence that the association of perceived severity with seropositivity differed significantly between genders (*p*-value of interaction = 0.3).

**Figure 6:**
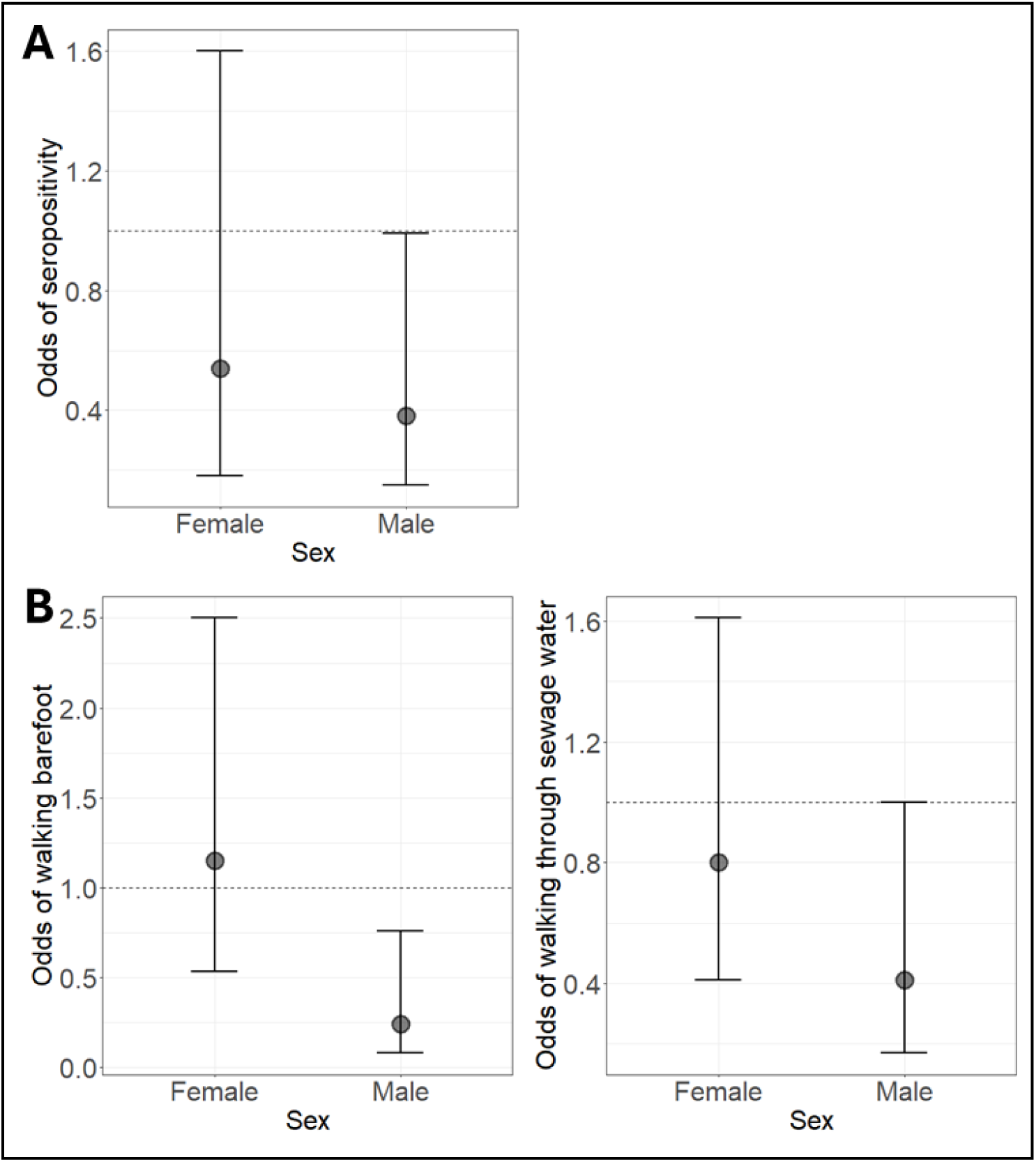
Total causal effects of perceived severity with A) seropositivity and B) high-risk behaviours. Odds ratios are shown for females or males who perceived leptospirosis as extremely serious compared to those of the same gender who perceived leptospirosis as less serious.

Among employed participants, high-risk occupations were associated with increased odds of seropositivity in the male-restricted model, but not in the combined or female-restricted models. In the male-restricted model, men who had high-risk occupations had 3.92 (95%CI 1.17, 13.2) times the odds of being seropositive than those that had other occupations. In the combined model, those in high-risk occupations had 1.60 (95%CI 0.72, 3.54) times the odds of being seropositive than those in other occupations. A conflicting estimate was found in the female-restricted model, as women in high-risk occupations had 0.92 (95%CI 0.23, 2.92) times the odds of being seropositive than those in other occupations, although the association was not significant at the 5% level. Despite the difference in point estimates, a test for interaction presented no evidence that the association of high-risk occupations with seropositivity differed significantly between genders (*p*-value of interaction = 0.07).

Other exposures, such as race, education, employment, and behaviours, were not associated with seropositivity in the combined or sex-disaggregated models.

**Table 2:**
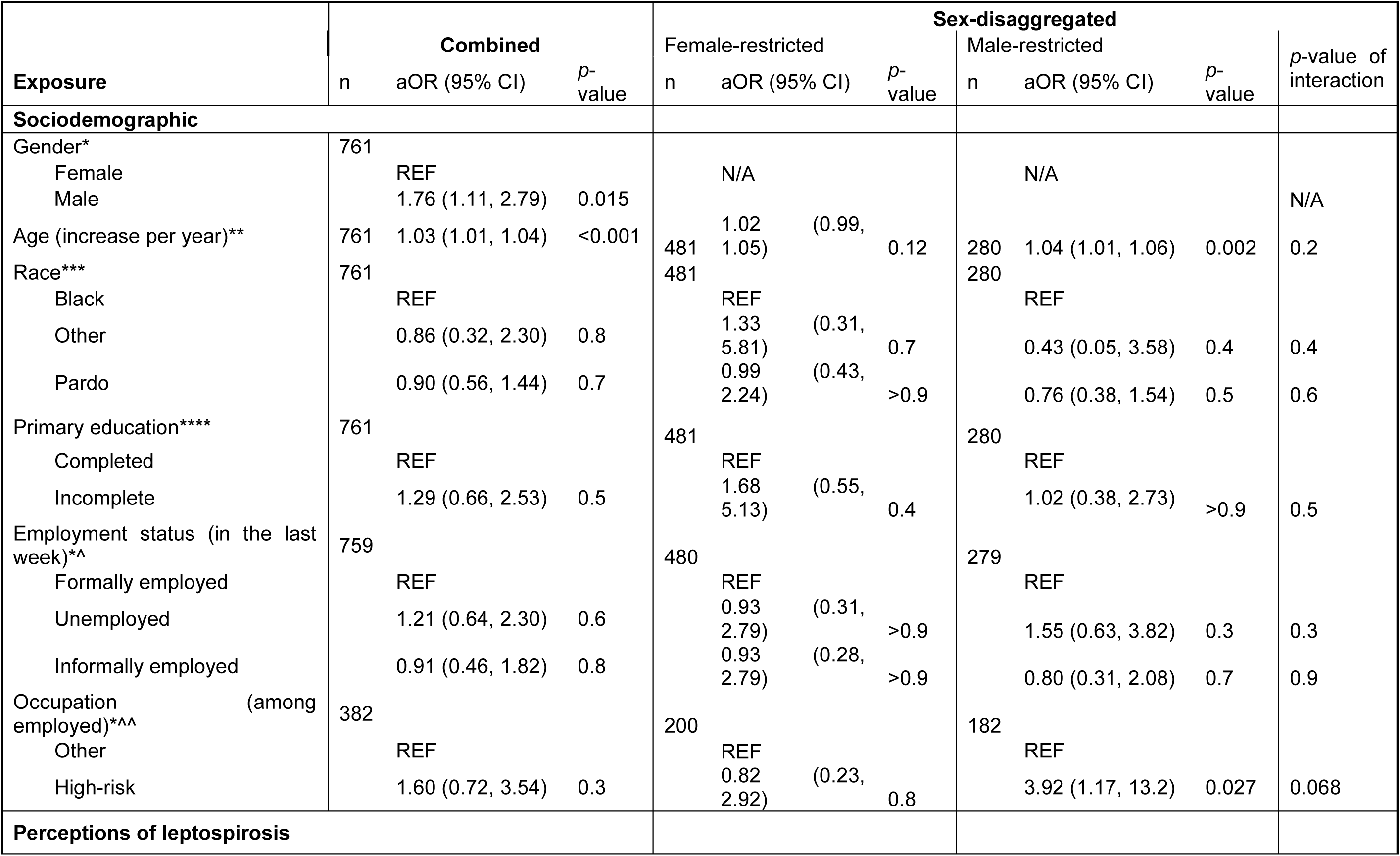
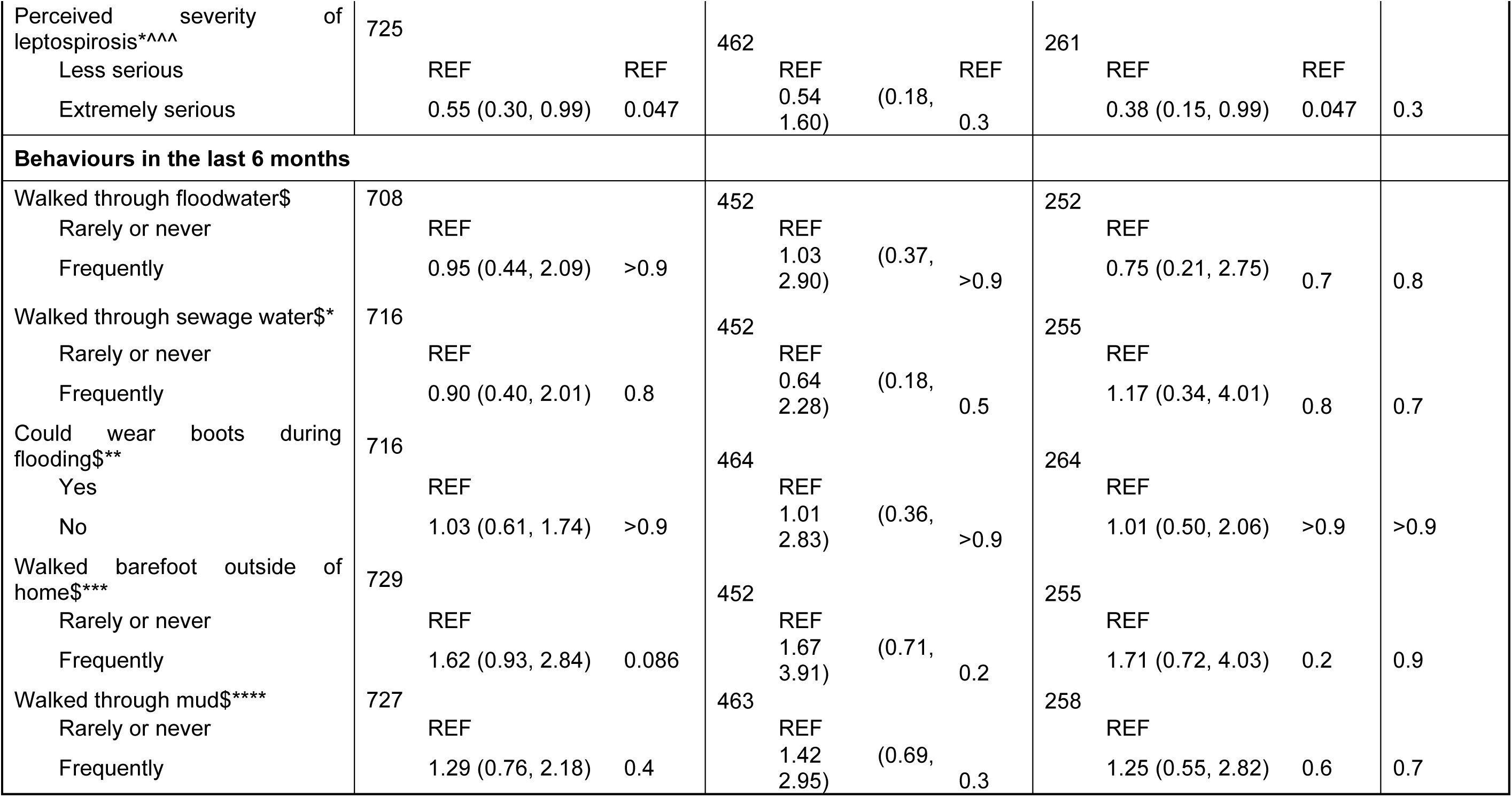
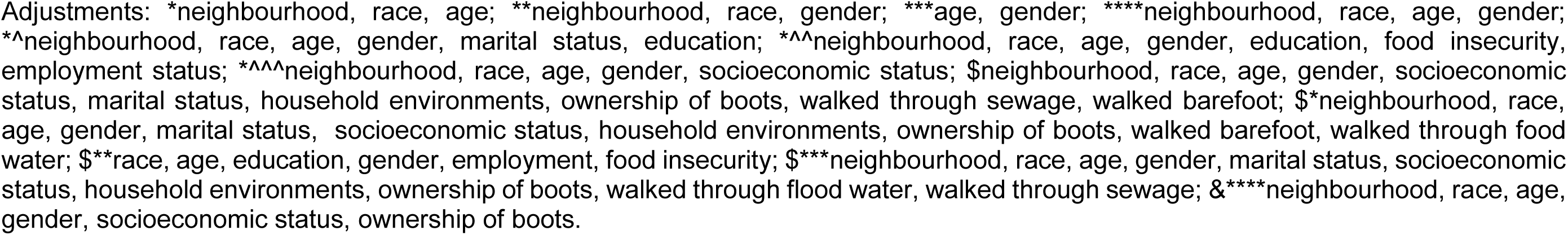
Total causal effect estimates for the effect of each exposure on seropositivity, shown for the combined and sex-disaggregated multivariable logistic regression models.

### Analysis 2: Perceived severity as a determinant of high-risk behaviours

To further investigate the finding that perceived severity of leptospirosis was associated with seropositivity and explore its role as a driver of seropositivity through behaviours (as illustrated in Figure 3), we examined the association between perceived severity and high-risk behaviours.

#### Descriptive

The sex-disaggregated descriptive analysis of the distribution of high-risk behaviours across perceived severity is included in Supplementary Information 3a. Notably, a smaller proportion of men who perceived leptospirosis as extremely serious reported walking barefoot outside of the home than those who perceived leptospirosis as less serious (21.4% vs. 41.2%).

#### Regression analyses

Uni- and multivariable analyses were used to estimate the total causal effect of perceived severity on the risk of performing high-risk behaviours. The univariable analysis is included in Supplementary Figure 3b. In each multivariable regression model, the functional form of age was identified in the GAM analysis (Supplementary Information 3c), resulting in age was modelled as a continuous variable or as a linear piecewise splines with knots at specific ages identified by the GAMs (Supplementary Information 3c). The full multivariable regression analysis exploring the association of perceived severity and behaviours is shown in Supplementary Figure 4c, with two key findings shown in Figure 6B.

Perceived severity was associated with lower odds of walking barefoot outside of the home in the male-restricted model but not in the female-restricted model (Figure 6B), or the combined model. In the male-restricted models, men who perceived leptospirosis as extremely serious had 0.24 (95%CI 0.08, 0.76) times the odds of walking barefoot outside of the home compared with men who perceived leptospirosis as less serious. In the female-restricted models, women who perceived leptospirosis as extremely serious had 1.15 (95%CI 0.53, 2.50) times the odds of walking barefoot than women who perceived leptospirosis as less serious, however the association was not significant at the conventional 5% significance level. A test for interaction provided further evidence to support that the association between perceived severity and walking barefoot differed significantly between genders (*p*-value of interaction = 0.007).

Perceived severity was also associated with reduced odds of walking through sewage water in the male-restricted model but not in the female restricted model (Figure 6B), or the combined model. In the male-restricted models, men who perceived leptospirosis as extremely serious had 0.41 (95%CI 0.17, 1.00) times the odds of walking through sewage water than men who perceived leptospirosis as less serious. In the female-restricted models, women who perceived leptospirosis as extremely serious had 0.80 (95%CI 0.41, 1.62) times the odds of walking through sewage water than women who perceived leptospirosis as less serious, however the association was not significant at the conventional 5% significance level. A test for interaction showed no evidence that the association of perceived severity with walking through sewage differed by gender (*p*-value of interaction = 0.3).

## Discussion

To the best of our knowledge, we present the first sex-disaggregated study to investigate gender-determined infection risk of *Leptospira*. By investigating sex-disaggregated associations of leptospirosis risk factors, we were able to identify sex-specific relationships that were not found in the combined analyses. First, we investigated sociodemographic, perception, and behavioural risk factors to understand which exposures contribute to *Leptospira* seroprevalences of 14.6% and 9.4% in men and women respectively. We found that leptospirosis was perceived as an extremely serious disease by most men and women, but that only men who perceived leptospirosis as extremely serious had a lower odds of being seropositive than men who perceived leptospirosis as less serious. Among employed participants, a similar proportion of men and women had high-risk occupations, but men in high-risk occupations had a much higher odds of being seropositive than those in other occupations. In the second analysis, we found that men who perceived leptospirosis as extremely serious had a lower odds of walking barefoot or walking through sewage water. None of these associations were found for female participants and there was evidence for gender as an effect modifier for the relationship between perceived severity and walking barefoot. Taken together, our results indicate the role of high-risk occupations, perceptions, and high-risk behaviours in driving differential risk by gender. Furthermore, they demonstrate the benefit of applying causal inference methodology to sex-disaggregated frameworks to disentangle complex gender-determined infection risks.

The finding that men who perceived leptospirosis as extremely serious were less likely to walk through sewage or walk barefoot outside of the home than those who did not, suggests that perceptions of disease severity may influence disease-specific preventive behaviours. This aligns with the Rational Model Theory (24). Walking through sewage water and walking barefoot are commonly cited risk factors for leptospirosis (4,47), particularly the latter with 11 out of 12 studies included in one systematic review identifying walking barefoot outside as a risk factor (16). These behaviours are particularly high-risk in this setting as the communities are built over open sewers that often overflow with heavy rain, dispersing leptospires that can enter through broken skin on the feet from the surrounding contaminated soil or water.

The fact that walking barefoot and walking through sewage water were the only behaviours associated with perceived severity could be explained by them being easier to moderate than the other measured behaviours, such as contact with flood water and mud, in this analysis. For residents of flood-prone marginalised communities with unpaved streets, these sources of environmental risk are often very challenging to avoid when moving within the community and exposure to them may be driven more strongly by household location than perceptions and attitudes to risk. Furthermore, the ability to wear boots during flooding is likely driven by socioeconomic circumstances, as was also found in the previous KAP study of leptospirosis risks (8). Conversely, participants may be able to avoid areas where sewers are known to overflow and avoid walking barefoot, and therefore these behaviours are more directly within a person’s control.

We found that sex was an effect modifier for the relationship between perceived severity and walking barefoot, with no evidence in the female-restricted model of a statistical association. We hypothesised that the lack of association between perceived severity and walking barefoot in the female-restricted model could be explained by women having much more limited access to boots than men (18% vs. 50%). For this reason, we tested to see whether the gender interaction could be explained by this difference in boot ownership but found no evidence of an interaction of boot ownership with perceived severity (*p*-value of interaction = 0.3). While the determinants underlying this gender interaction effect remain uncertain, this finding highlights the importance of conducting sex-disaggregated or gender-interaction analyses to identify gender-specific relationships that are obscured in combined model.

Despite perceived severity and seropositivity being associated in men and the point estimate for women also being in the same direction, none of the high-risk behaviours analysed were associated with *Leptospira* seropositivity. This may be indicative of the relative contributions of two infection risks: 1) periods of intense flooding exposure in and around the household, and 2) low-intensity exposure to soil and mud. It is likely that our study population had a homogeneous and high environmental risk from flooding events because the study areas were relatively small and were selected to include households located in the lowest sections of the communities, where the risk of leptospirosis is known to be highest (15,30). Therefore, environmental exposure due to heavy flooding may ultimately mean that the effect of preventive behaviours, such as wearing shoes, is limited. To better understand this causal pathway, future studies should consider studying areas with greater variation in environmental risk to identify behaviours with a substantial impact on individual risk.

The absence of statistical associations observed in female-restricted models suggests that the pathways leading to exposure in women may not be fully captured. Given that male gender is commonly cited as a risk factor in leptospirosis studies in Salvador and globally, the measured exposures may be biased toward male gender norms (4,12,13). To address this, potential exposures aligned with female cultural norms in Salvador should be incorporated to distinguish infection risks in women. For instance, women in Salvador are traditionally associated with limited mobility in the environment and engage in domestic roles within the home (23). Consequently, exposure to *Leptospira* may occur in the household environment during these activities. Future serosurveys should thus consider expanding the scope of questions to encompass behavioural exposures that may be more prevalent among women.

Due to the neglected status of leptospirosis and the broader gap in sex-disaggregated CSIDs research, limited gender-specific data hinders our understanding of infection risks and behavioural exposures in women (10,22,48,49). To address this gap, qualitative focus group discussions segregated by gender may help to shape hypotheses surrounding exposures in women, which could subsequently be measured and tested in serosurveys (50). This mixed-methods approach allows for open-ended questioning to capture unanticipated information and has been of benefit in South Africa to reveal gender-based exposures to leptospirosis (51,52). Identifying and measuring exposures more prevalent in women could help tailor interventions to both men and women, thus improving our understanding and control of leptospirosis transmission.

We found a linear increase in the risk of *Leptospira* seropositivity with age. While it is conventionally characterized that younger adult age groups are at higher risk (12,13), our findings are consistent with previous studies in the city of Salvador that have also identified a linear trend with age (4,15). This may also be partially attributable to the persistence of antibodies, which can remain detectable for several years following exposure and reflects a limitation of this study (53). The persistence of antibodies over several years presents additional challenges as behaviours were self-reported during the previous six months as a measure of exposure risk (53). This, and challenges accurately measuring behaviour using self-reported questions, are limitations of our study and may have resulted in measurement error.

Additionally, while serostatus is a practical and easily interpretable measure of *Leptospira* exposure given the commonly subclinical and non-specific febrile symptoms, it may not capture gender inequalities in disease burden, particularly as there is evidence that progression of leptospirosis to SPHS is more common in women than men (21). Therefore, inclusion of broader disease burden metrics that encompass both prevalence and outcomes may better address gender disparities in leptospirosis research.

Finally, while our study provides a benefit for beginning to understand the causal pathways of gender-related risks, the use of binary gender categories of men and women presents a limitation. Future studies should consider broader data collection methods that accommodate diverse gender identities and explore the intersectional effects of other social determinants of health, such as race and socioeconomic status.

## Conclusion

As the first sex-disaggregated study investigating *Leptospira* infection risks, our findings suggest an important but complex role for perceptions of disease severity and high-risk behaviours that appear to shape gender-determined infection risk of leptospirosis. This underscores the critical need for sex-disaggregated frameworks in leptospirosis research, but also in the study of other zoonotic CSIDs affected by similar flooding hazards in urban environments, such as Toxoplasma gondii.

More broadly, gender-sensitive approaches to climate change and CSID research are important to understand the intersectional biological, social, and environmental drivers that contribute to health outcomes. Future studies incorporating gender intersectional analyses can support the development of equitable and climate-resilient interventions that account for the differential risks experienced by men, women, boys, girls, and other gender identities.

## Data Availability

All data produced in the present study are available upon reasonable request to the authors

https://dagitty.net/dags.html?id=XxPTXytr#

## Acknowledgements

We thank the residents and community leaders of Nova Sussuarana, Arenoso, Calabetão, and Jardim Santo Inácio/Mata Escura, for their support and participation in this study. We would also like to thank the community driving group (GIC-Grupo Impulsor Comunitario) in the communities Nova Sussuarana and Arenoso for their support. This work was supported by the Wellcome Trust (218987/Z/19/Z), Oswaldo Cruz Foundation and Secretariat of Health Surveillance, Brazilian Ministry of Health. ED was supported by a London School of Hygiene and Tropical Medicine MSc Travel Scholarship. MTE was supported by a Reckitts Global Hygiene Institute (RGHI) fellowship. AK was supported by the NIHR (R01 AI121207, U01 AI088752, R01 TW009504). FC was supported by NIH/NIAID (F31 AI114245, R01 AI052473, U01 AI088752, R01 TW009504, R25 TW009338), Fundação de Amparo à Pesquisa do Estado da Bahia (FAPES-45 B/JCB0020/2016), and the Wellcome Trust (102330/Z/13/Z).

## Supplementary Information

**Supplementary Information 1:**
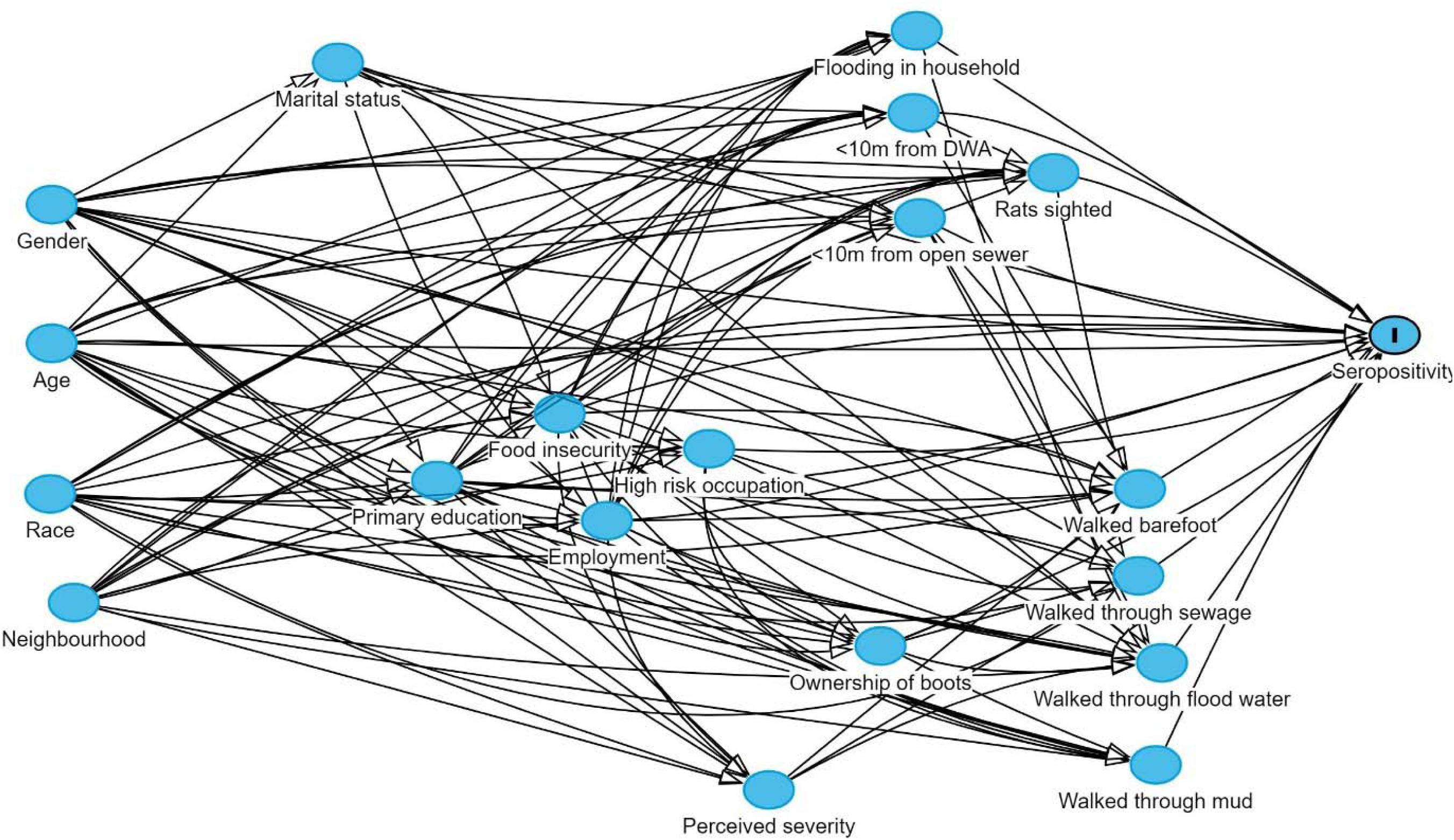
Full version of the DAG used in analysis, also available at https://dagitty.net/dags.html?id=XxPTXytr<u>#</u>. DWA: Domestic waste accumulation

**Supplementary Information 2a:** Sex-disaggregated descriptive analysis of seroprevalence across risk-factors.

|  | Females |  | Males |  |
| --- | --- | --- | --- | --- |
|  | Total | Seroprevalence<br>n (%) | Total | Seroprevalence<br>n (%) |
| Sociodemographic |  |  |  |  |
| Age group (years) | 481 |  | 280 |  |
| 18-30 |  | 8 (6.5) |  | 5 (6.4) |
| 31-45 |  | 15 (8.9) |  | 13 (12.9) |
| 46-60 |  | 12 (10.0) |  | 13 (18.3) |
| >60 |  | 10 (14.7) |  | 10 (33.3) |
| Race | 481 |  | 280 |  |
| Black |  | 22 (9.1) |  | 23 (15.9) |
| Other |  | 4 (12.1) |  | 1 (6.7) |
| Pardo |  | 19 (9.2) |  | 17 (14.2) |
| Completion of primary education | 481 |  | 280 |  |
| Yes |  | 6 (5.9) |  | 6 (10.2) |
| No |  | 39 (10.3) |  | 35 (15.8) |
| Employment status (as of previous week) | 480 |  | 279 |  |
| Unemployed |  | 27 (9.8) |  | 19 (20.2) |
| Formal |  | 6 (9.2) |  | 12 (12.8) |
| Informal |  | 12 (8.6) |  | 10 (11.0) |
| Occupation | 480 |  | 279 |  |
| High-risk |  | 4 (8.2) |  | 10 (21.7) |
| Other |  | 14 (9.0) |  | 12 (8.6) |
| Unemployed |  | 27 (9.8) |  | 19 (20.2) |
| Perceptions of leptospirosis |  |  |  |  |
| Perceived severity of leptospirosis | 474 |  | 272 |  |
| Less serious |  | 9 (14.3) |  | 9 (26.5) |
| Extremely serious |  | 36 (8.8) |  | 31 (13.0) |
| Behaviours (in last 6 months) |  |  |  |  |
| Walked through flood water | 474 |  | 272 |  |
| Rarely |  | 35 (10.1) |  | 32 (15.1) |
| Frequently |  | 10 (7.9) |  | 9 (15.0) |
| Walked through sewage water | 474 |  | 273 |  |
| Rarely |  | 38 (10.2) |  | 31 (14.6) |
| Frequently |  | 7 (7.0) |  | 10 (16.4) |
| Could wear boots during flooding | 475 |  | 273 |  |
| No |  | 37 (9.5) |  | 21 (15.3) |
| Yes |  | 8 (9.3) |  | 20 (14.7) |
| Walked barefoot | 474 |  | 273 |  |
| Rarely |  | 33 (9.0) |  | 29 (14.0) |
| Frequently |  | 12 (11.1) |  | 12 (18.2) |
| Walked through mud | 475 |  | 273 |  |
| Rarely |  | 32 (9.0) |  | 29 (14.8) |
| Frequently |  | 13 (10.8) |  | 12 (15.6) |

**Supplementary Information 2b:** Sex-disaggregated univariable logistic regression analysis.

| Exposure | Combined |  |  | Sex-disaggregated |  |  | Sex-disaggregated |  |  |
| --- | --- | --- | --- | --- | --- | --- | --- | --- | --- |
|  | n | OR (95% CI) | p-value | Female-restricted<br>n | OR (95% CI) | p-value | Male-restricted<br>n | OR (95% CI) | p-value |
| **Sociodemographics** |  |  |  |  |  |  |  |  |  |
| Gender | 761 |  |  |  |  |  |  |  |  |
| Female |  | REF | REF |  | N/A |  |  | N/A |  |
| Male |  | 1.66 (1.06, 2.61) | 0.027 |  |  |  |  |  |  |
| Age (increase per year) | 761 | 1.02 (1.01, 1.04) | 0.001 | 481 | 1.02 (1.00, 1.05) | 0.12 | 280 | 1.04 (1.01, 1.06) | 0.002 |
| Race | 761 |  |  | 481 |  |  | 280 |  |  |
| Black |  | REF | REF |  | REF | REF |  | REF | REF |
| Other |  | 0.88 (0.33, 2.34) |  |  | 1.45 (0.34, 6.14) | 0.6 |  | 0.38 (0.05, 3.02) | 0.4 |
| Pardo |  | 0.94 (0.59, 1.49) | 0.8 |  | 1.00 (0.45, 2.22) | >0.9 |  | 0.88 (0.44, 1.73) | 0.7 |
| Primary education | 761 |  |  | 481 |  |  | 280 |  |  |
| Completed |  | REF | REF |  | REF | REF |  | REF | REF |
| Incomplete |  | 1.75 (0.92, 3.30) | 0.086 |  | 2.01 (0.68, 5.96) | 0.2 |  | 1.66 (0.66, 4.16) | 0.3 |
| Employment status (in the last week) | 759 |  |  | 480 |  |  | 279 |  |  |
| Formally employed |  | REF | REF |  | REF | REF |  | REF | REF |
| Unemployed |  | 1.11 (0.62, 1.99) | 0.7 |  | 1.06 (0.33, 3.39) | >0.9 |  | 1.73 (0.79, 3.81) | 0.2 |
| Informally employed |  | 0.83 (0.43, 1.60) | 0.6 |  | 0.86 (0.23, 3.15) | 0.8 |  | 0.84 (0.35, 2.06) | 0.7 |
| Occupation (among employed) | 389 |  |  | 204 |  |  | 185 |  |  |
| Other |  | REF | REF |  | REF | REF |  | REF | REF |
| High-risk |  | 1.78 (0.89, 3.57) | 0.1 |  | 0.90 (0.24, 2.64) | 0.9 |  | 3.50 (0.41, 29.6) | 0.3 |
| **Perceptions of leptospirosis** |  |  |  |  |  |  |  |  |  |
| Perceived severity of leptospirosis | 746 |  |  | 474 |  |  | 272 |  |  |
| Less serious |  | REF | REF |  | REF | REF |  | REF | REF |
| Extremely serious |  | 0.51 (0.29, 0.89) | 0.019 |  | 0.54 (0.20, 1.47) | 0.2 |  | 0.42 (0.18, 0.97) | 0.043 |
| **Behaviours in the last 6 months** |  |  |  |  |  |  |  |  |  |
| Walked through floodwater | 746 |  |  | 474 |  |  | 272 |  |  |
| Rarely or never |  | REF | REF |  | REF | REF |  | REF | REF |
| Frequently |  | 0.84 (0.49, 1.43) | 0.5 |  | 0.71 (0.26, 1.89) | 0.5 |  | 0.99 (0.45, 2.21) | >0.9 |
| Walked through sewage water | 747 |  |  | 474 |  |  | 273 |  |  |
| Rarely or never |  | REF | REF |  | REF | REF |  | REF | REF |
| Frequently |  | 0.88 (0.50, 1.55) | 0.7 |  | 0.55 (0.17, 1.78) | 0.3 |  | 1.14 (0.53, 2.49) | 0.7 |
| Could wear boots during flooding | 748 |  |  | 475 |  |  | 273 |  |  |
| Yes |  | REF | REF |  | REF | REF |  | REF | REF |
| No |  | 0.86 (0.53, 1.39) | 0.5 |  | 1.02 (0.36, 2.90) | >0.9 |  | 1.05 (0.54, 2.04) | 0.9 |
| Walked barefoot outside of home | 747 |  |  | 474 |  |  | 273 |  |  |
| Rarely or never |  | REF | REF |  | REF | REF |  | REF | REF |
| Frequently |  | 1.32 (0.80, 2.19) | 0.3 |  | 1.41 (0.54, 3.68) | 0.5 |  | 1.36 (0.65, 2.85) | 0.4 |
| Walked through mud | 748 |  |  | 475 |  |  | 273 |  |  |
| Rarely or never |  | REF | REF |  | REF | REF |  | REF | REF |
| Frequently |  | 1.17 (0.71, 1.92) | 0.5 |  | 1.31 (0.53, 3.27) | 0.6 |  | 1.06 (0.51, 2.21) | 0.9 |

**Supplementary Information 2c:**
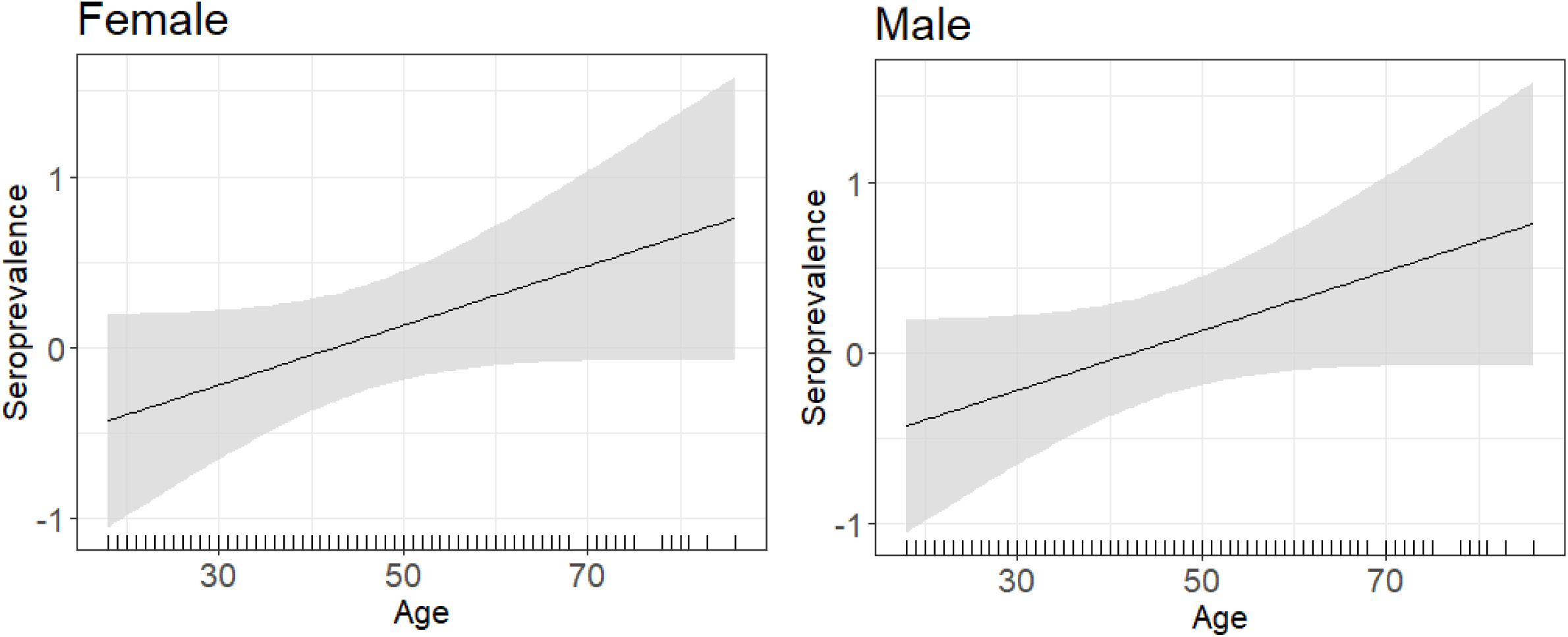
GAMs of age with seroprevalence response variable. GAMs were built using univariable models, and shaded area corresponds to 95%CI.

**Supplementary Information 3a:**
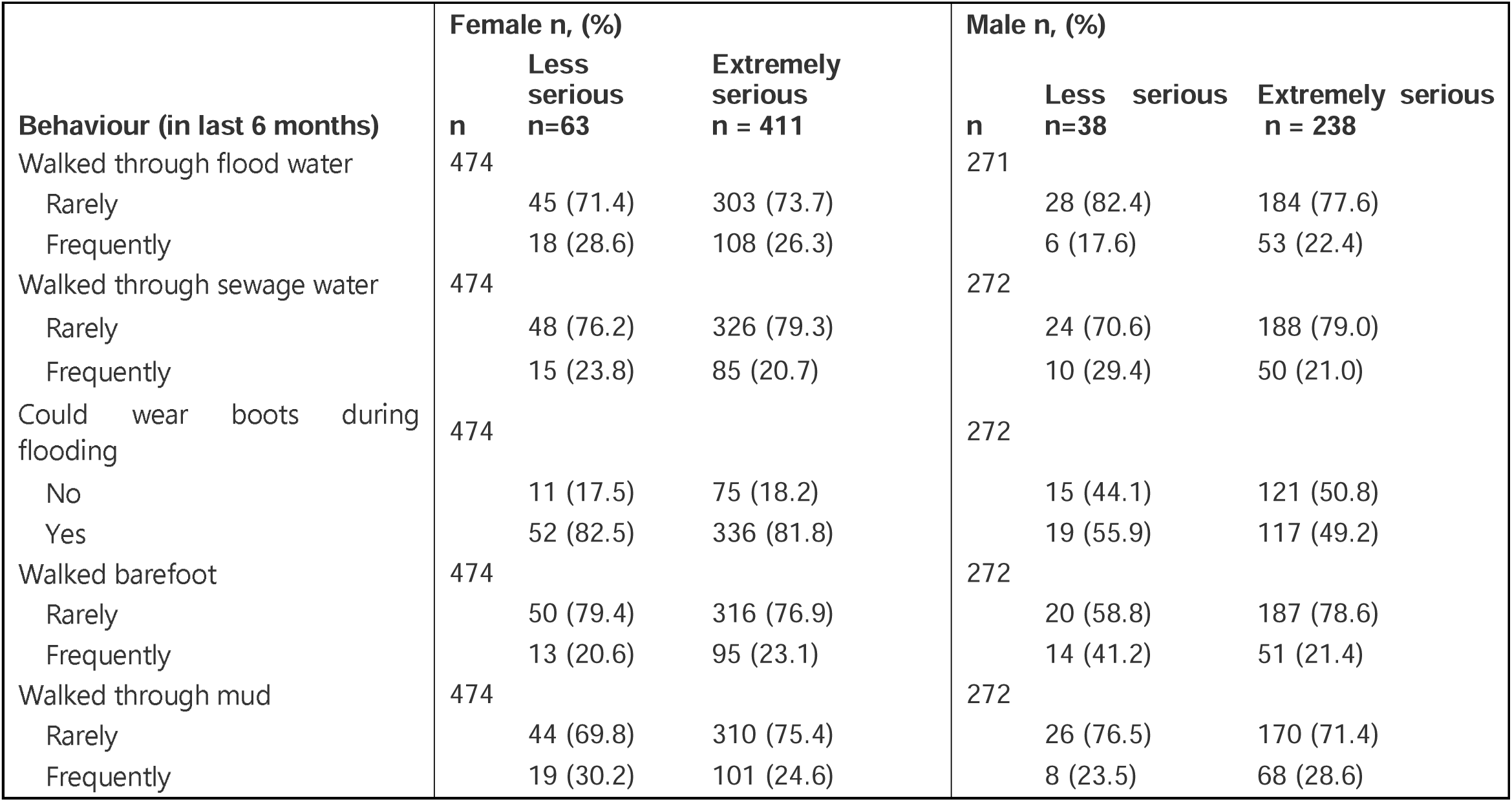
Descriptive analysis of prevalence of behaviours across perceived severity.

**Supplementary Information 3b:**
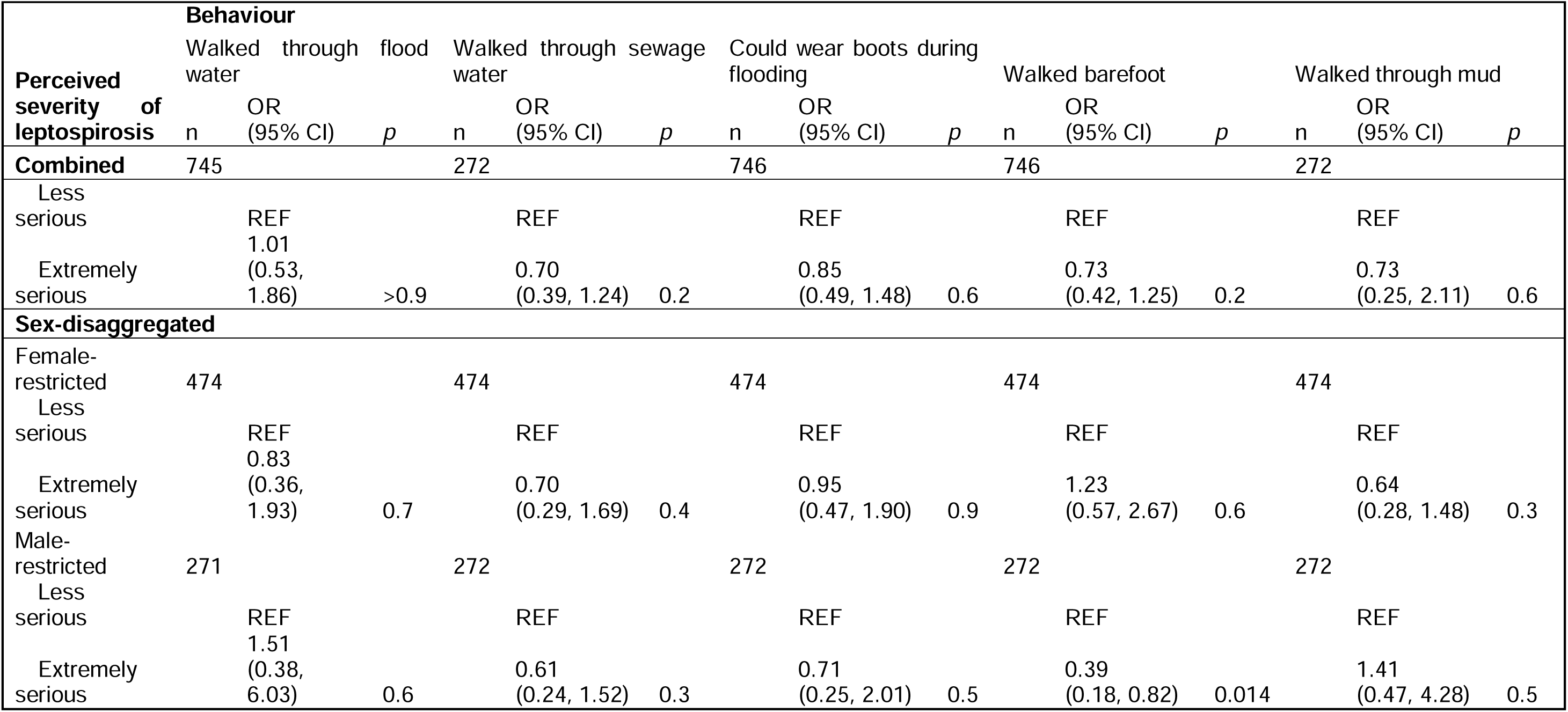
Sex-disaggregated univariable logistic regression analysis of the association of perceived severity with high-risk behaviours.

**Supplementary 3c:**
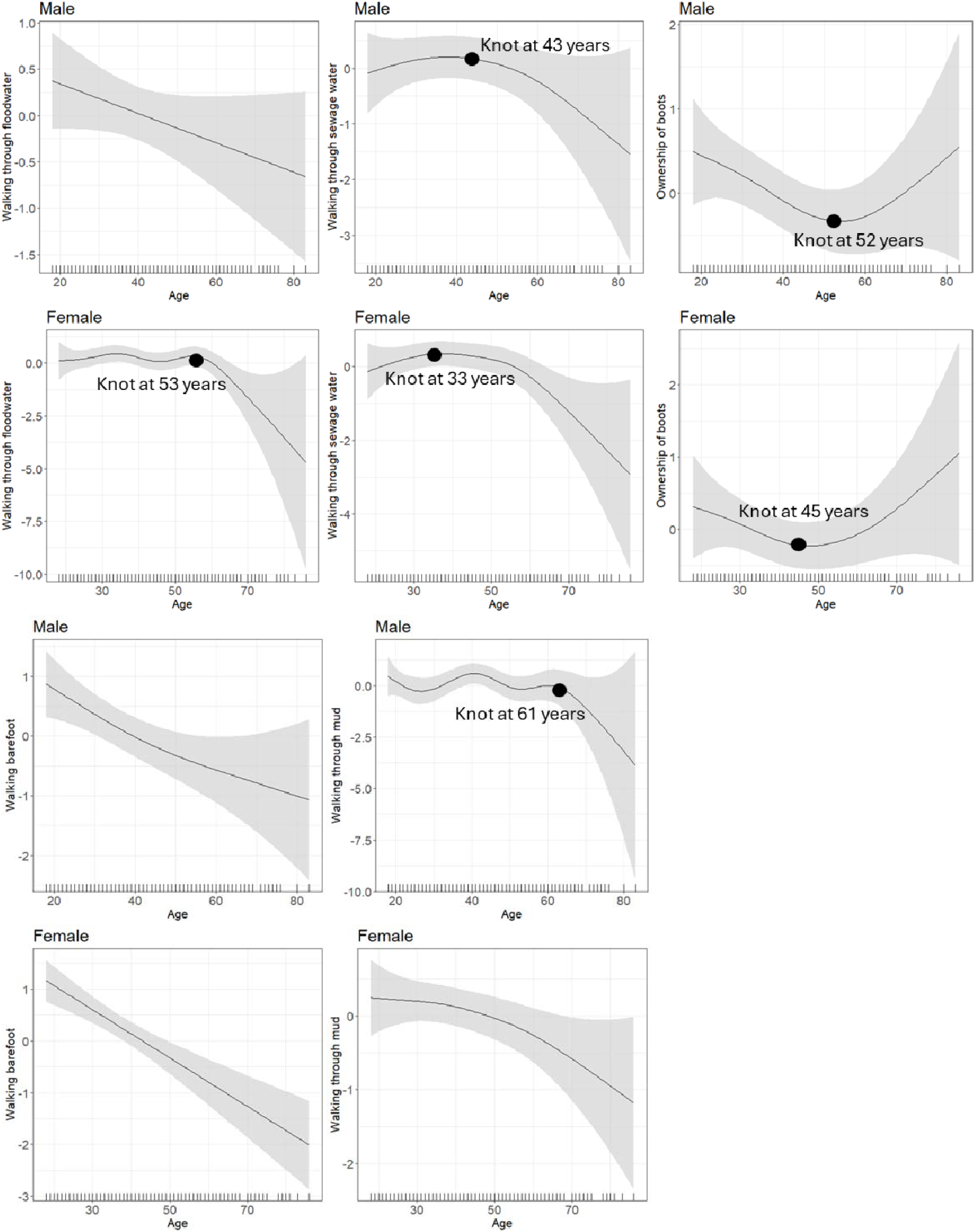
GAMs of age with high-risk behaviour response variables, and position of knots used to model non-linear relationships. GAMs were built using univariable models, and shaded area corresponds to 95%CI.

**Supplementary Information 3d:**
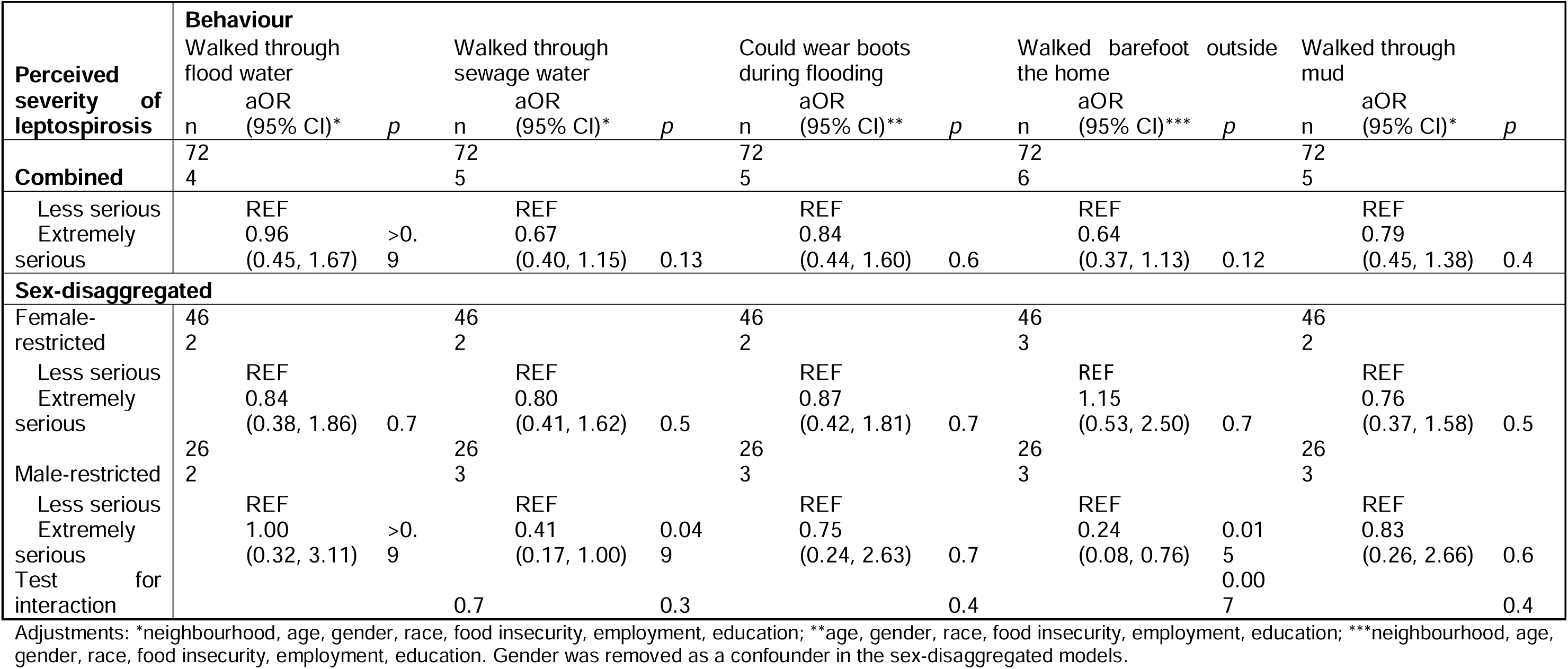
Total causal effect estimates for the effect of perceived severity on the risk of performing high-risk behaviours, shown for the combined and sex-disaggregated multivariable logistic regression models.

